# Association of appendectomy with decreased risk of cardiovascular events in elderly Australian patients

**DOI:** 10.1101/2025.02.28.25323127

**Authors:** Courtney P. Judkins, Yutang Wang, Alexander Bobik, Christopher G. Sobey, Grant R. Drummond

**Affiliations:** Centre for Cardiovascular Biology and Disease Research, La Trobe Institute for Molecular Sciences, La Trobe University, Bundoora, Victoria, 3086, Australia; Department of Microbiology, Anatomy, Physiology & Pharmacology, School of Agriculture, Biomedicine & Environment, La Trobe University, Bundoora, Victoria, 3086, Australia; Discipline of Life Science, Institute of Innovation, Science and Sustainability, Federation University Australia, Ballarat, Victoria, 3350, Australia; Baker Heart and Diabetes Institute, Melbourne, Victoria, 3004, Australia

**Keywords:** appendix, appendectomy, cardiovascular disease, myocardial infarction, stroke

## Abstract

**Background:** The appendix is thought to contribute to gut and systemic immune system function, and thus could influence cardiovascular health and disease progression. However, it is unclear if there is an association between appendectomy and cardiovascular risk. This study aimed to estimate the strength of any such association in 978,095 Australian hospital patients aged ≥60 years, among which 5,130 underwent appendectomy.

**Methods:** The association of appendectomy with cardiovascular events (myocardial infarction, angina, stroke and transient ischemic attack) was analysed using binary logistic regression adjusting for age, sex, cardiovascular risk factors, and gastrointestinal disorders.

**Results:** Appendectomy was inversely associated with overall cardiovascular risk (odds ratio [OR], 0.73; 95% confidence interval [CI], 0.66-0.81; P<0.001), myocardial infarction (OR, 0.71; 95% CI, 0.63-0.80; P<0.001) and stroke (OR, 0.77; 95% CI, 0.66-0.90; P=0.001) after adjustment for all the test confounders. This inverse association existed in both sexes and remained when hernia surgery was used as a control instead of patients without appendectomy.

**Conclusions:** This study found that appendectomy was associated with >27% lower multivariate-adjusted risk for overall cardiovascular events, myocardial infarction, or stroke, in patients aged 60 years and over. Our results provide additional support for the current clinical practice of removing the appendix in the elderly when appendicitis is suspected.

## INTRODUCTION

The appendix vermiformis, or appendix, is a tubular pouch with an average length of 9 cm^1^ that branches off the cecum near the ileocecal valve^2^. The appendix serves as a secondary lymphoid organ, containing mucosa-associated lymphoid tissue^3^, and is involved in lymphocyte maturation and IgA antibody production^2,4^.

Acute appendicitis is inflammation of the appendix and a common cause of abdominal pain^5^. The annual incidence of appendicitis is approximately 100 cases per 100,000 in the adult population worldwide^6,7^. Appendectomy, or surgical removal of the appendix, is the gold standard for treating acute appendicitis^8–10^, and is performed in approximately 60% of patients with appendicitis^11^. It is estimated that the lifetime risk of appendicitis is 16% and the lifetime risk of appendectomy is 10%^11^.

Recent evidence suggests that appendectomy may have additional impacts on health beyond its primary purpose of resolving acute appendicitis. Some studies suggest that appendectomy may have a negative impact on health, as it is associated with an increased risk for irritable bowel syndrome^12^, Crohn’s disease^13^ and mood and anxiety disorders^14^. However, others suggest there may be health benefits associated with removal of the appendix. For example, appendectomy was shown to be associated with a reduced risk for ulcerative colitis^15,16^ and some neurodegenerative diseases including Parkinson’s disease^17^ and amyotrophic lateral sclerosis^18^.

There is also evidence that appendectomy may influence cardiovascular health. Indeed, there have been two studies that investigated the association between appendectomy and cardiovascular events^3,19^. Janszky et al^3^ evaluated the association of appendectomy with acute myocardial infarction using Swedish national register data including 54,449 participants who underwent appendectomy before 20 years of age and 272,213 control subjects^3^. The results showed that appendectomy was associated with a 33% higher risk for acute myocardial infarction. Chen et al^19^ similarly investigated the association between appendectomy and ischemic heart disease in 5,413 adult patients who underwent appendectomy (mean age: 39.4 years) and 16,239 control subjects (mean age: 39.2 years) from the Longitudinal Health Insurance Database 2000 in Taiwan. The authors also found that appendectomy was associated with a 54% higher risk of ischemic heart disease^19^.

It is noteworthy that in these earlier studies the vast majority of patients were young (<40 years), both at the time of their surgery and even after the follow up period (3-20 years). It is well established that major adverse cardiovascular events are relatively rare in patients prior to middle age. Furthermore, the predominant causes of these events in younger people are likely to be different from those in older individuals. Thus, whereas in younger individuals factors such as congenital heart defects, coronary artery spasm, familial hypercholesterolemia and cerebral aneurysm account for a significant proportion of cardiovascular events, in older people, long-term exposure to risk factors such as hypertension, high cholesterol, diabetes and smoking – many of which are associated with underlying inflammation – is likely to play a more important role. Therefore, we reasoned that the effects of appendectomy on cardiovascular health may also differ in young versus old patients. Specifically, we hypothesised that removal of the appendix and any associated inflammatory burden may be more likely to have beneficial effects on cardiovascular health in older patients. Therefore, to test this hypothesis, we conducted a population level study on 978,095 Australian hospital patients to compare the risk of cardiovascular events occurring in individuals who received an appendectomy at ≥60 years versus that in age-matched non-appendectomised patients.

## METHODS

### Study participants

Study participants were provided by the Data Custodian of the Centre for Victorian Data Linkage, the Department of Health and Human Services of Victoria^20^, with underlying data derived from all hospitals in Victoria, Australia^21^. From 1 July 2000 to 30 June 2020, 8,064,624 patients were recorded to be admitted to Victorian hospitals in the dataset. A total of 6,273,406 patients who were aged <60 years at first admission were excluded. Among the remaining 1,791,218 patients, 18,803 had appendectomy surgery (the appendectomy group) and 1,772,415 did not (the control group). In addition, patients were excluded if they received cancer treatment or had suffered from a cardiovascular event prior to or within 6 months of appendectomy surgery or first admission. The final analysis included a total of 978,095 patients aged ≥60 years, 5,130 of whom underwent appendectomy surgery (**Figure 1**).

**Figure 1:**
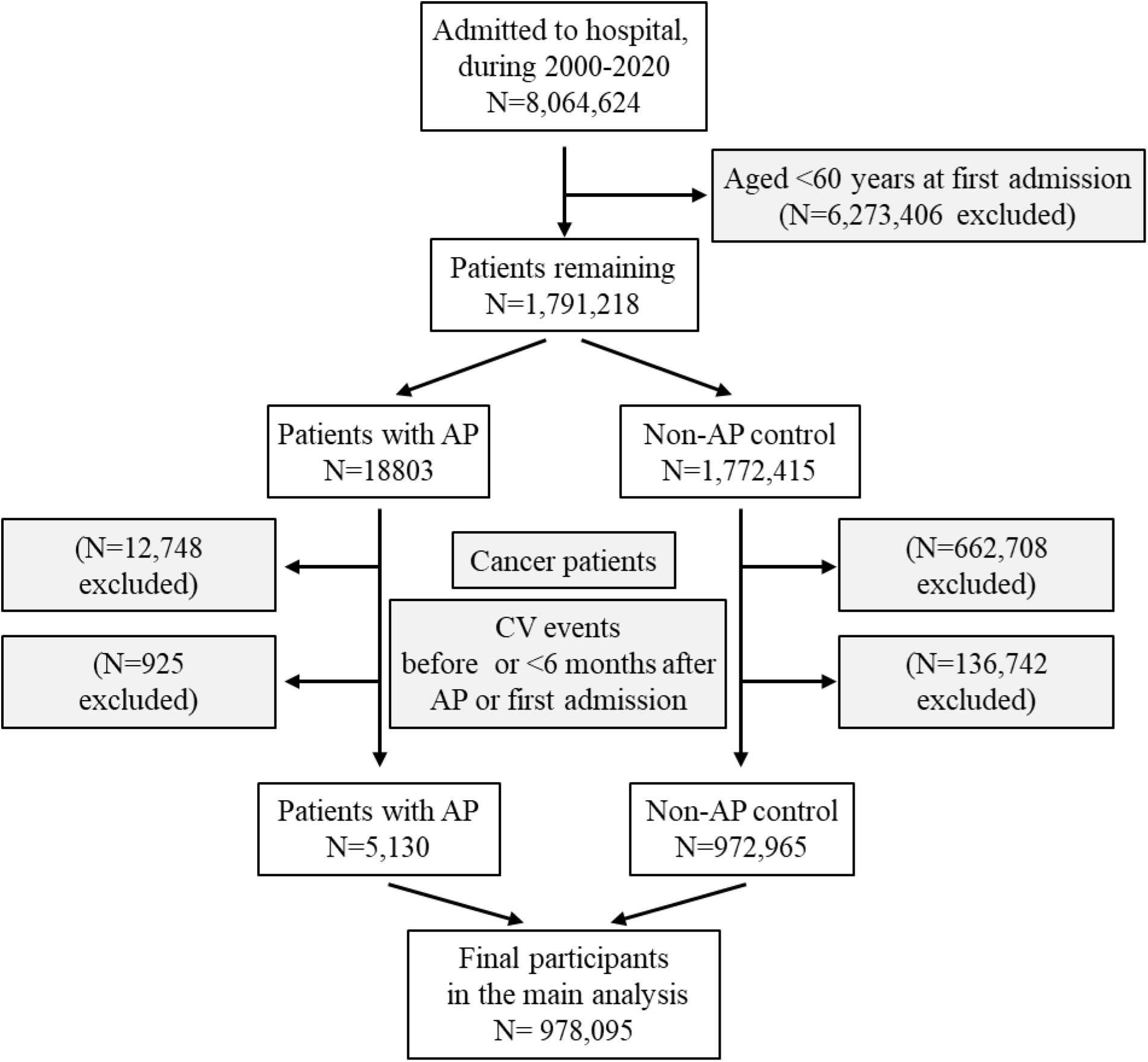
Flow diagram of the study participants. AP, appendectomy.

### Ethical considerations

This study was approved by La Trobe University Human Research Ethics Committee (approval number, HEC20380). All procedures were performed following the guidelines of the Declaration of Helsinki. Patient information was anonymized and de-identified prior to analysis and written informed consent was waived by the La Trobe University Human Research Ethics Committee.

### Appendectomy

Appendectomy surgery was identified according to the Australian Classification of Health Interventions codes 3057100 and 3057200.

### Cardiovascular events

Cardiovascular events in the current study included myocardial infarction (I21, I22, I23, I24, I25), angina (I200), stroke (G46, I60, I61, I62, I63, I64, I65, I66, I67), and transient ischaemic attack (G450-453, G458-459), which were diagnosed according to the International Classification of Diseases, 10th Revision (Supplementary Table 1).

### Covariates

Confounding covariates included age (continuous), sex (male or female), obesity (yes or no), hypertension (yes or no), diabetes (yes or no), sleep apnoea (yes or no), atrial fibrillation and cardiac arrhythmia (yes or no), irritable bowel syndrome (yes or no), ulcerative colitis (yes or no), and Crohn’s disease (yes or no). Obesity (E66 or U781), hypertension (I10, I11, I12, I13, or I15), diabetes (E10, E11, E13, or E14), sleep apnoea (G473, G474, G478, or G479), COPD (U832), atrial fibrillation and cardiac arrhythmia (I48, I49), irritable bowel syndrome (K58), ulcerative colitis (K51 or U842), and Crohn’s disease (K50 or U841) were diagnosed according to the International Classification of Diseases, 10th Revision^22^ (Supplementary Table 1).

### Statistical Analyses

Data were presented as mean and standard deviation for continuous variables, or number and percentage for categorical variables^23^. Differences in age between two groups were analysed using Student’s t-test^24^. Differences among categorical variables were analysed using Fisher’s exact test^25,26^. The associations of appendectomy with cardiovascular risks were analysed by binary logistic regression with cardiovascular events as the outcome variable, adjusted for age, sex, obesity, diabetes, sleep apnoea, atrial fibrillation and arrhythmia, irritable bowel syndrome, ulcerative colitis, and Crohn’s disease.

Further analyses were conducted to assess the association of appendectomy with cardiovascular events using patients undergoing hernia surgery as the control instead of all the hospital patients naïve to appendectomy. Of the 972,965 control patients included in the main analysis, 26,866 patients were identified as hernia surgery patients and were treated as the hernia surgery control group. Within the appendectomy surgery group in the main analysis, 246 patients were identified to have also undergone hernia surgery and were excluded, resulting in a final number of 4,884 appendectomy surgery patients for analysis (Supplementary Figure 1). Hernia surgery was identified according to the Australian Classification of Health Interventions codes 3060902 and 3061402._The null hypothesis was rejected using a two-tailed P-value of <0.05. All analyses were performed using Stata/SE 18 (Standard Edition 18.0, Stata Corporation, College Station, Texas, USA).

## RESULTS

### General characteristics of the participants

The study included 978,095 patients aged ≥60 years, among whom 5,130 underwent appendectomy surgery. Compared with the control patients who did not undergo appendectomy surgery, those who underwent appendectomy surgery were younger, more obese, and more likely to have comorbidities including hypertension, sleep apnoea, atrial fibrillation and arrhythmia, irritable bowel syndrome, ulcerative colitis, and Crohn’s disease (**Table 1**)

**Table 1:** Characteristics of participants, appendectomy versus non-appendectomy control patients.

|  | **Appendectomy** | **Control** | **Overall** | **P value** |
| --- | --- | --- | --- | --- |
| **Variable** |  |  |  |  |
| Sample size | 5130 | 972965 | 978095 | NA |
| Age, y, mean (SD) | 68.9 (7.8) | 73.8 (9.6) | 73.8 (9.6) | <0.001 |
| Male, N, (%) | 2343 (45.7) | 425505 (43.7) | 427848 (43.7) | 0.005 |
| Hypertension, N (%) | 960 (18.7) | 168604 (17.3) | 169564 (17.3) | 0.010 |
| Obesity, N (%) | 306 (6.0) | 25790 (2.6) | 26096 (2.7) | <0.001 |
| Diabetes, N (%) | 743 (14.5) | 147261 (15.1) | 148004 (15.1) | 0.197 |
| Sleep apnoea, N (%) | 250 (4.9) | 21539 (2.2) | 21789 (2.2) | <0.001 |
| Atrial Fibrillation and Arrhythmia, N (%) | 742 (14.5) | 125842 (12.9) | 126584 (12.9) | 0.001 |
| Irritable bowel syndrome, N (%) | 53 (1.0) | 2623 (0.3) | 2676 (0.3) | <0.001 |
| Ulcerative colitis, N (%) | 36 (0.7) | 3248 (0.3) | 3284 (0.3) | <0.001 |
| Crohn's disease, N (%) | 43 (0.8) | 1622 (0.2) | 1665 (0.2) | <0.001 |

N, number; SD, standard deviation.

### Major cardiovascular events in appendectomy versus control patients

Compared with controls, patients who had undergone appendectomy experienced fewer cardiovascular events (10.0% versus 11.3%, P=0.005), fewer strokes (3.4% versus 4.1%, P=0.012), and fewer myocardial infarctions (6.3% versus 7.1%, P=0.024, **Table 2**).

**Table 2:** Major cardiovascular events in appendectomy versus non-appendectomy control patients.

|  | Appendectomy | Control | Overall | P value |
| --- | --- | --- | --- | --- |
| Cardiovascular event, N (%) | 515 (10.0) | 109613 (11.3) | 110128 (11.3) | 0.005 |
| Cerebral vascular event, N (%) | 221 (4.3) | 48372 (5.0) | 48593 (5.0) | 0.028 |
| All strokes, N (%) | 173 (3.4) | 39485 (4.1) | 39658 (4.0) | 0.012 |
| Haemorrhagic stroke, N (%) | 37 (0.7) | 9060 (0.9) | 9097 (0.9) | 0.125 |
| Ischemic stroke, N (%) | 104 (2.0) | 19304 (2.0) | 19408 (2.0) | 0.802 |
| TIA, N (%) | 68 (1.3) | 13416 (1.4) | 13484 (1.4) | 0.810 |
| Myocardial event, N (%) | 342 (6.7) | 73731 (7.6) | 74073 (7.6) | 0.014 |
| Myocardial infarction, N (%) | 322 (6.3) | 68928 (7.1) | 69250 (7.1) | 0.024 |
| Angina, N (%) | 73 (1.4) | 16649 (1.7) | 16722 (1.7) | 0.117 |

N, number; TIA, transient ischemic attack.
Cardiovascular events include myocardial infarction, angina, stroke, and TIA.
Cerebral vascular events include stroke and TIA.
Myocardial events include myocardial infarction and angina.

### Inverse association of appendectomy with cardiovascular events

Appendectomy was inversely associated with cardiovascular events after adjustment for age and sex (odds ratio [OR], 0.90; 95% confidence interval [CI], 0.82-0.98; P=0.020; Table 3). The inverse association remained significant after further adjustment for all the tested comorbidities (OR, 0.73; 95% CI, 0.66-0.81; P<0.001; Model 3; Table 3). Further analyses showed that appendectomy was inversely associated with myocardial infarction (OR, 0.71; 95% CI, 0.63-0.80; P<0.001; Model 3; Table 3) and stroke (OR, 0.77; 95% CI, 0.66-0.90; P=0.001; Model 3; Table 3) after adjustment for all the test confounders.

**Table 3:** Odds ratio of cardiovascular events associated with appendectomy, using patients who did not undergo appendectomy as control.

|  | Cardiovascular event |  | Myocardial infarction |  | All stroke |  |
| --- | --- | --- | --- | --- | --- | --- |
|  | OR (95% CI) | P value | OR (95% CI) | P value | OR (95% CI) | P value |
| All patients |  |  |  |  |  |  |
| Model 1 | 0.90 (0.82-0.98) | 0.020 | 0.85 (0.76-0.96) | 0.007 | 0.90 (0.78-1.05) | 0.188 |
| Model 2 | 0.74 (0.66-0.82) | <0.001 | 0.72 (0.63-0.81) | <0.001 | 0.78 (0.66-0.91) | 0.002 |
| Model 3 | 0.73 (0.66-0.81) | <0.001 | 0.71 (0.63-0.80) | <0.001 | 0.77 (0.66-0.90) | 0.001 |
| Male patients |  |  |  |  |  |  |
| Model 1 | 1.00 (0.89-1.13) | 0.965 | 0.98 (0.85-1.13) | 0.800 | 0.93 (0.75-1.16) | 0.536 |
| Model 2 | 0.79 (0.69-0.91) | 0.001 | 0.79 (0.67-0.93) | 0.004 | 0.76 (0.61-0.96) | 0.022 |
| Model 3 | 0.78 (0.68-0.90) | 0.001 | 0.78 (0.67-0.92) | 0.003 | 0.76 (0.60-0.96) | 0.021 |
| Female patients |  |  |  |  |  |  |
| Model 1 | 0.79 (0.69-0.91) | 0.001 | 0.70 (0.58-0.84) | <0.001 | 0.88 (0.71-1.08) | 0.223 |
| Model 2 | 0.68 (0.58-0.79) | <0.001 | 0.62 (0.51-0.76) | <0.001 | 0.79 (0.64-0.98) | 0.036 |
| Model 3 | 0.67 (0.58-0.78) | <0.001 | 0.61 (0.51-0.75) | <0.001 | 0.79 (0.63-0.98) | 0.031 |

CI, confidence interval; OD: odds ratio.
Model 1: adjusted for age and sex
Model 2: adjusted for age, sex, hypertension, obesity, diabetes, sleep apnoea, atrial flutter and fibrillation
Model 3: adjusted for factors in Model 2 plus irritable bowel syndrome, Crohn's disease and ulcerative colitis.

Sub-analyses were conducted in each sex sub-group. The results showed that the inverse association of appendectomy with cardiovascular events remained in male and female participants (**Table 3**).

### Inverse association of appendectomy procedure with cardiovascular events, when hernia surgery was used as a control

Further analyses were conducted to rule out the possibility that the observed inverse association between appendectomy and cardiovascular events was simply derived from general surgery procedures rather than being specific for appendix removal. In these analyses, appendectomy was compared with hernia surgery.

Compared to the patients who underwent hernia surgery, those who underwent appendectomy were younger, more obese, and more likely to have comorbidities including hypertension, diabetes, sleep apnoea, irritable bowel syndrome, and Crohn’s disease (**Table 4**).

**Table 4:** Characteristics of participants in patients undergoing appendectomy or hernia surgery.

| Variable | Appendectomy | Hernia surgery | Overall | P value |
| --- | --- | --- | --- | --- |
| Sample size | 4884 | 26866 | 31750 | NA |
| Male, N, (%) | 2137 (43.8) | 23315 (86.8) | 70.6 (8.1) | <0.001 |
| Age, y, mean (SD) | 68.8 (7.8) | 70.96 (8.1) | 25452 (80.2) | <0.001 |
| Hypertension | 920 (18.8) | 4466 (16.6) | 5386 (17.0) | <0.001 |
| Obesity | 297 (6.1) | 659 (2.4) | 956 (3.0) | <0.001 |
| Diabetes | 709 (14.5) | 3293 (12.3) | 4002 (12.6) | <0.001 |
| Sleep apnoea | 240 (4.9) | 1007 (3.7) | 1247 (3.9) | <0.001 |
| Atrial Fibrillation and Arrhythmia | 695 (14.2) | 3910 (14.5) | 4605 (14.5) | 0.566 |
| Irritable bowel syndrome | 52 (1.1) | 121 (0.4) | 173 (0.5) | <0.001 |
| Ulcerative colitis | 34 (0.7) | 156 (0.6) | 190 (0.6) | 0.315 |
| Crohn's disease | 42 (0.9) | 94 (0.3) | 136 (0.4) | <0.001 |

Compared with patients that underwent hernia surgery, appendectomy patients experienced fewer cardiovascular events (9.8% versus 14.1%, P<0.001), including fewer strokes (3.4% versus 4.5%, P<0.001), and fewer myocardial infarctions (6.1% versus 9.6%, P<0.001, **Table 5**).

**Table 5:**
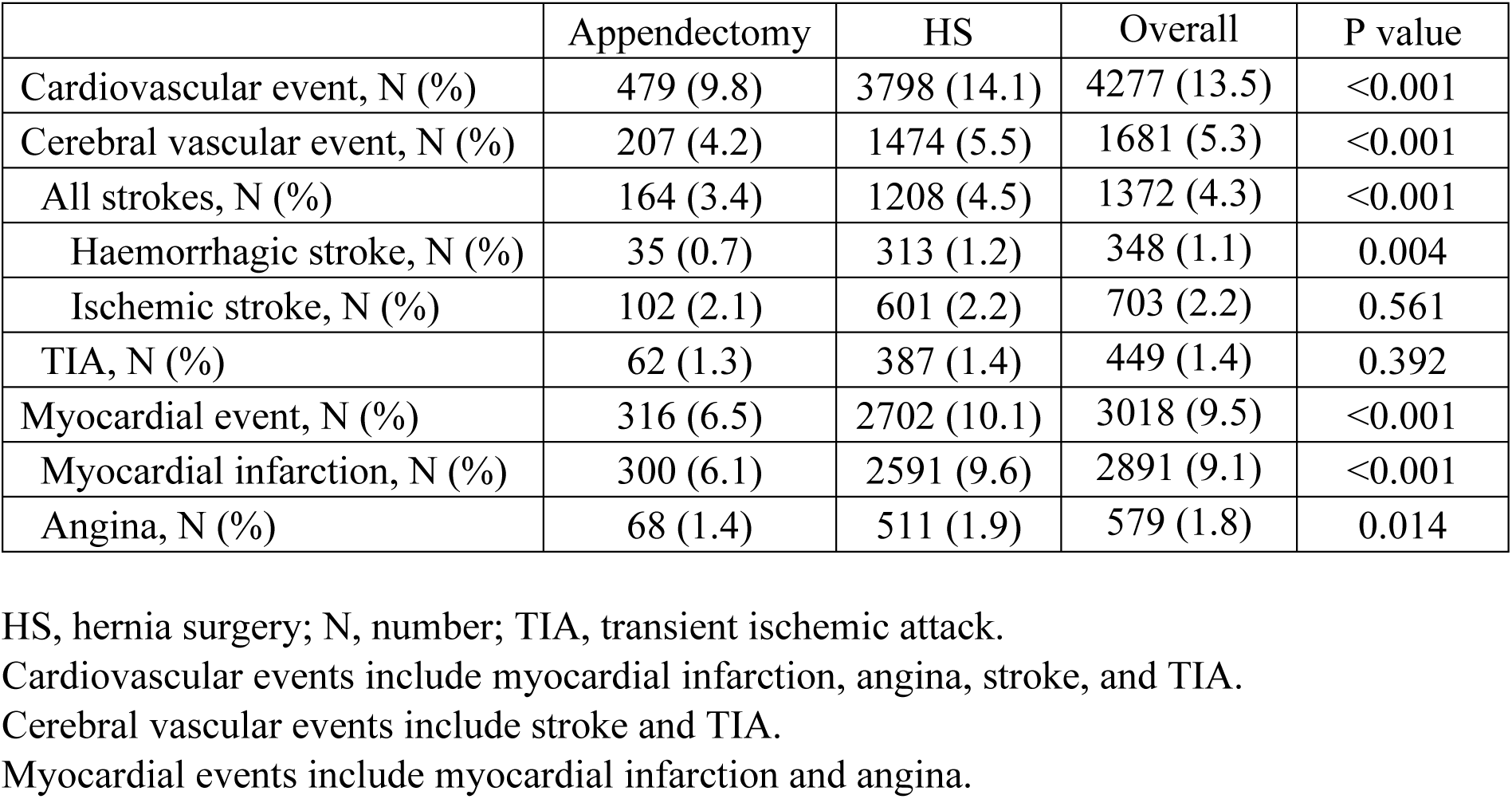
Major cardiovascular events in patients undergoing appendectomy or hernia surgery.

|  | Appendectomy | HS | Overall | P value |
| --- | --- | --- | --- | --- |
| Cardiovascular event, N (%) | 479 (9.8) | 3798 (14.1) | 4277 (13.5) | <0.001 |
| Cerebral vascular event, N (%) | 207 (4.2) | 1474 (5.5) | 1681 (5.3) | <0.001 |
| All strokes, N (%) | 164 (3.4) | 1208 (4.5) | 1372 (4.3) | <0.001 |
| Haemorrhagic stroke, N (%) | 35 (0.7) | 313 (1.2) | 348 (1.1) | 0.004 |
| Ischemic stroke, N (%) | 102 (2.1) | 601 (2.2) | 703 (2.2) | 0.561 |
| TIA, N (%) | 62 (1.3) | 387 (1.4) | 449 (1.4) | 0.392 |
| Myocardial event, N (%) | 316 (6.5) | 2702 (10.1) | 3018 (9.5) | <0.001 |
| Myocardial infarction, N (%) | 300 (6.1) | 2591 (9.6) | 2891 (9.1) | <0.001 |
| Angina, N (%) | 68 (1.4) | 511 (1.9) | 579 (1.8) | 0.014 |

HS, hernia surgery; N, number; TIA, transient ischemic attack.
Cardiovascular events include myocardial infarction, angina, stroke, and TIA.
Cerebral vascular events include stroke and TIA.
Myocardial events include myocardial infarction and angina.

Compared with hernia surgery, appendectomy was inversely associated with overall cardiovascular events (OR, 0.66; 95% CI, 0.59-0.75; P<0.001; **Table 6**) after adjustment for all the tested confounders. Further analyses showed that, compared with hernia surgery, appendectomy was inversely associated with myocardial infarction (OR, 0.65; 95% CI, 0.57- 0.75; P<0.001; **Table 6**) and stroke (OR, 0.77; 95% CI, 0.64-0.92; P=0.004; **Table 6**) after adjustment for all the tested confounders.

**Table 6:** Odds ratio of cardiovascular events associated with appendectomy, using hernia surgery as a control.

|  | Cardiovascular event |  | Myocardial infarction |  | All stroke |  |
| --- | --- | --- | --- | --- | --- | --- |
|  | OR (95% CI) | P value | OR (95% CI) | P value | OR (95% CI) | P value |
| All patients |  |  |  |  |  |  |
| Model 1 | 0.80 (0.72-0.89) | <0.001 | 0.77 (0.68-0.88) | <0.001 | 0.86 (0.72-1.03) | 0.095 |
| Model 2 | 0.67 (0.59-0.75) | <0.001 | 0.65 (0.57-0.75) | <0.001 | 0.77 (0.64-0.92) | 0.005 |
| Model 3 | 0.66 (0.59-0.75) | <0.001 | 0.65 (0.57-0.75) | <0.001 | 0.77 (0.64-0.92) | 0.004 |
| Male patients |  |  |  |  |  |  |
| Model 1 | 0.86 (0.75-0.99) | 0.034 | 0.87 (0.74-1.02) | 0.081 | 0.86 (0.68-1.09) | 0.220 |
| Model 2 | 0.66 (0.57-0.77) | <0.001 | 0.68 (0.57-0.81) | <0.001 | 0.72 (0.56-0.92) | 0.009 |
| Model 3 | 0.66 (0.57-0.77) | <0.001 | 0.68 (0.57-0.81) | <0.001 | 0.72 (0.56-0.92) | 0.009 |
| Female patients |  |  |  |  |  |  |
| Model 1 | 0.77 (0.64-0.92) | 0.003 | 0.66 (0.52-0.83) | <0.001 | 0.91 (0.69-1.19) | 0.490 |
| Model 2 | 0.71 (0.59-0.85) | <0.001 | 0.61 (0.48-0.78) | <0.001 | 0.86 (0.65-1.14) | 0.287 |
| Model 3 | 0.69 (0.57-0.84) | <0.001 | 0.61 (0.48-0.77) | <0.001 | 0.85 (0.64-1.13) | 0.273 |

CI, confidence interval; OD: odds ratio.
Model 1: adjusted for age and sex
Model 2: adjusted for age, sex, hypertension, obesity, diabetes, sleep apnoea, atrial flutter and fibrillation
Model 3: adjusted for factors in Model 2 plus irritable bowel syndrome, Crohn's disease and ulcerative colitis.

Sub-analyses were conducted in each sex sub-group. The results showed that the inverse association of appendectomy with cardiovascular events remained in male and female participants (**Table 6**).

## DISCUSSION

This study found that appendectomy was associated with a 27-29% lower multivariate-adjusted risk for either total cardiovascular events, myocardial infarction, or stroke, in Australian patients aged 60 years and over. This inverse association existed in both males and females and remained when hernia surgery was used as a control instead of hospital patients without appendectomy.

The appendix is a secondary lymphoid organ and may play a role in protecting against pathogens.^2,3^ The appendix has abundant microfold (M) cells^27^, which are characterised by their ability to take up antigens from the appendicular lumen via endocytosis, phagocytosis and transcytosis and deliver them to antigen-presenting cells, which then interact with B and T lymphocytes in local lymphoid follicles^28^. Thus, the appendix plays an important role in lymphocyte maturation. In addition, the appendix may play a role in the production of IgA antibodies^2^. Thus, it is plausible that appendectomy may alter the body’s immune capacity, and in turn impact on the susceptibility/resistance to certain immune-related diseases. Myocardial infarction and stroke are two of the major causes of mortality and morbidity globally, especially in individuals beyond middle age. Given that immune cell activation and inflammation are established to contribute to many of the risk factors for cardiovascular events, such as hypertension, diabetes and atherosclerosis^29–31^, we postulated that appendectomy might reduce cardiovascular disease progression and thus lower the incidence of major cardiovascular events. We further reasoned that any beneficial effects of appendectomy would be most apparent in older individuals in whom the incidence of cardiovascular events is high and more likely to be attributable to cumulative exposure to systemic inflammation associated with the aforementioned risk factors. Hence, we limited the present study to patients aged ≥60 years. Indeed, we found that appendectomy after age 60 was associated with a markedly reduced risk of major cardiovascular events, including myocardial infarction and stroke.

Our findings thus contrast those of two previous studies in which it was shown that appendectomy is associated with an elevated risk of myocardial infarction^3,19^. Janszky et al^3^ reported that appendectomy, performed before 20 years of age, was associated with a 33% higher risk for myocardial infarction in participants in a Swedish national register^3^. Likewise, Chen et al^19^ reported that appendectomy was associated with a 54% higher risk of ischemic heart disease in people (mean age = 39 years) from a Taiwan health insurance database. When we used the current dataset to examine the impact of appendectomy prior to age 20 on subsequent cardiovascular events over a 20-year follow, we too saw a trend towards a positive association, although this failed to reach statistical significance (Supplementary Table 2).

The precise mechanism(s) for the differing effects of appendectomy on cardiovascular events in young versus older patients is currently unknown. One possibility for why appendectomy at younger ages has detrimental effects could relate to its influence on diabetes. Lee et al showed that appendectomy prior to middle age was associated with an increased risk of type 2 diabetes, whereas in patients who received appendectomy at later ages, this association was absent^32^.

Although Chen et al. adjusted for diabetes (as well as hypertension and hyperlipidemia) in their study^19^, no such adjustments were made by Janszky et al^3^. Hence, in this latter study, the potential contribution of diabetes to the increased risk of myocardial infarction post appendectomy cannot be ruled out.

Another mechanism by which appendectomy at earlier ages may increase the risk of cardiovascular events relates to its crucial role in adaptive immunity and protection against pathogens. As mentioned above, the appendix is a major site of antigen presentation and hence it plays a crucial role in educating T and B cells and protecting the host against invading pathogens. However, with aging, T and B cell function steadily declines. Thus, appendectomy at an early age is likely to have a greater impact on the ability of the host to fight infections than it would at later ages. Given that infections are known risk factors for cardiovascular events^33,34^, this could explain why appendectomy increases the risk of myocardial infarction in young but not in older individuals.

Finally, a notable difference between the current and previous studies is the inclusion herein of a 6-month washout period to minimise the likelihood of capturing post-surgical thrombotic events in our estimation of total cardiovascular events. According to the American Society of Hematology the risk of thrombotic events, such as venous thromboembolism, is significantly elevated for up to 12 weeks post-operatively^35,36^. We also took the additional step of using age-matched patients who underwent hernia surgery – for whom the risk of post-operative thrombotic events is likely to be similar – as an alternative control group and still observed the marked degree of protection from cardiovascular events following appendectomy in patients over 60.

The previous discussion focusses on the appendectomy surgery as the potential driver of the altered cardiovascular risk in young versus older patients. However, it is possible that the appendicitis episode itself (i.e. rather than the surgical procedure) is responsible for the increased risk observed in younger individuals via its impact on the production of pathogenic autoantibodies. Antibody-producing plasma cells are long lived and can produce pathogenic autoantibodies for many years after an event, retaining memory of the event and influencing future, apparently unrelated events^37^. Autoantibodies are produced following acute appendicitis including multiple IgM and IgG anti-lipopolysaccharide antibodies, the latter being generated by long lived plasma cells generated within germinal centres of the appendix. ^38^ Postmortem studies indicate that germinal centres within the appendix are reduced with age and largely undetectable around 60 years of age^39^, suggesting that pathogenic IgG autoantibodies are unlikely to be produced in significant quantities in elderly humans. Anti-lipopolysaccharide autoantibodies avidly bind lipopolysaccharides (LPS) and resultant LPS- immune complexes are potent inducers of inflammation^40,41^. Given that circulating immune complexes containing chlamydial lipopolysaccharide accumulate in infarcted human hearts, it is plausible that they may contribute to cardiac pathology via pro-inflammatory mechanisms in younger but not elderly appendicitis patients^42^.

Regardless of the mechanisms underlying the cardioprotection observed in patients who receive appendectomy post 60 years of age, our findings highlight appendectomy as a novel factor to consider when calculating patients’ overall cardiovascular risk profile. They may also influence clinicians’ attitudes toward negative appendectomy, i.e. the removal of a macroscopically normal appendix (which is later confirmed to be histologically normal) based on the presence of acute clinical symptoms that strongly suggest appendicitis. Negative appendectomy is relatively common, accounting for approximately 9-34% of all appendectomy cases^43–45^. Two surveys suggested that approximately 60% of surgeons in America and Europe favour the decision to remove a macroscopically normal appendix when appendicitis is suspected^46,47^. Our findings may provide further justification for such decisions in older patients, especially those identified to be at high risk for cardiovascular events.

The strengths of this study include its large sample size and adjustment for a swathe of key clinical confounders. Nevertheless, our study also has limitations. First, there is the possibility of procedure/disease misclassification during the coding process. However, appendectomy and cardiovascular outcomes in our study were identified by well-audited and standard methods of diagnostic coding. A previous study has shown that the accuracy of the coding quality for diagnosis in Victorian public hospitals is high, with a kappa of 0.91 between coding auditors and hospital coders.^48^ Second, the clinical parameters captured in the present dataset are non-continuous, that is, there is no information about the severity/level of the various comorbidities (e.g. hypertension, high cholesterol, diabetes, etc.) and thus the extent to which they are likely to have contributed to cardiovascular events in a given patient. Finally, information about certain patient characteristics that could potentially affect the incidence of cardiovascular events are missing from our dataset including dietary confounders (e.g. low fibre intake^49^) and medications.

In conclusion, our study highlights the potential cardioprotective effects of appendectomy in older adults, contrasting with previous findings in younger populations. These results underscore the importance of considering age and immune function when evaluating the long-term impacts of appendectomy on cardiovascular health. Future research should aim to elucidate the mechanisms behind these age-dependent differences to better inform clinical decisions and patient care.

## Data Availability

We used de-identified hospital records provided by the Victorian Department of Human Health Services. These data cannot be made publicly available.

## Funding

This research was funded by the National Health and Medical Research Council of Australia (grant numbers GNT2003752 and GNT2003156).

## Author contributions

Conceptualization: C.G.S. and G.R.D.; data analysis: C.P.J. and Y.W.; writing - original draft preparation: Y.W. and C.P.J.; writing - review and editing: Y.W., C.P.J., A.B., C.G.S. and G.R.D.; and funding acquisition: C.G.S. and G.R.D.

## Data availability

The datasets are available from the corresponding author upon approval from the Data Custodian of the Centre for Victorian Data Linkage.

## Competing interests

The authors declare no competing interests.

---

**Supplementary Figure 1:**
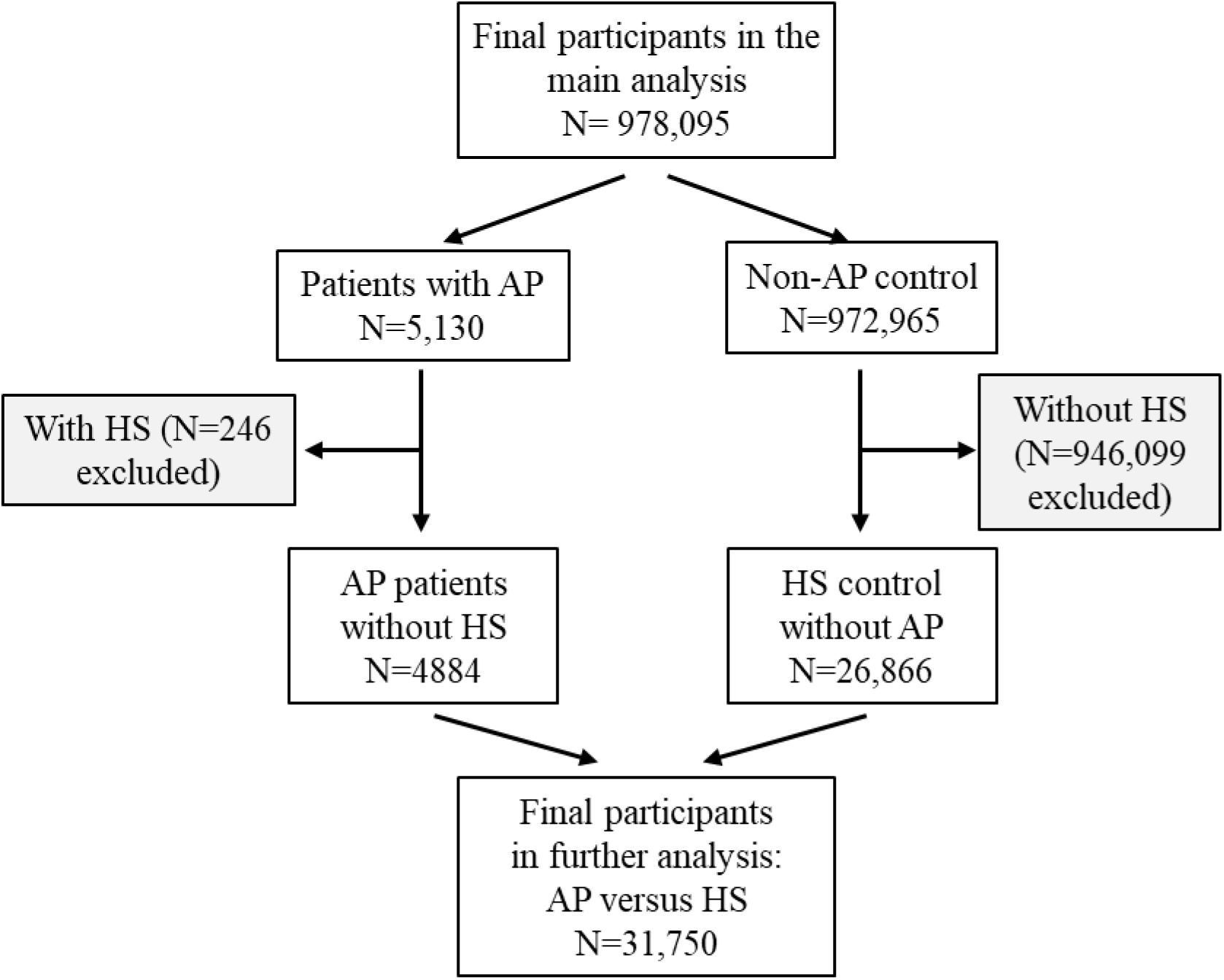
Flow diagram of the study participants in the further analysis of appendectomy versus hernia surgery. AP, appendectomy; HS, hernia surgery.

**Supplementary Table 1:** ICD10 codes for cardiovascular events and comorbidities.

| Cardiovascular events | ICD10 codes |
| --- | --- |
| Myocardial infarction | I21, I22, I23, I24, I25 |
| Angina | I200 |
| Stroke | G46, I60, I61, I62, I63, I64, I65, I66, I67 |
| Haemorrhagic stroke | I60, I61, I62 |
| Ischemic stroke | I63 |
| Transient ischaemic attack | G450-453, G458-459 |
| Comorbidities |  |
| Hypertension | I10, I11, I12, I13, I15 |
| Obesity | E66, U781 |
| Diabetes | E10, E11, E13, E14 |
| Sleep apnea | G473, G474, G478, G479 |
| Atrial fibrillation and arrhythmia | I48, I49 |
| Irritable bowel syndrome | K58 |
| Ulcerative colitis | K51, U842 |
| Crohn's disease | K50, U841 |

ICD10: the International Classification of Diseases, 10th Revision

**Supplementary Table 2:** Odds ratio of cardiovascular events associated with appendectomy in patients who underwent appendectomy before 20 years of age (N=46,471), using patients who did not undergo appendectomy as control (N=2,092,908)

|  | Cardiovascular event |  | Myocardial infarction |  | All stroke |  |
| --- | --- | --- | --- | --- | --- | --- |
|  | OR (95% CI) | P value | OR (95% CI) | P value | OR (95% CI) | P value |
| All patients |  |  |  |  |  |  |
| Model 1 | 0.96 (0.74-1.26) | 0.794 | 1.25 (0.76-2.05) | 0.378 | 1.02 (0.73-1.40) | 0.921 |
| Model 2 | 0.93 (0.71-1.22) | 0.600 | 1.27 (0.77-2.10) | 0.342 | 0.98 (0.70-1.35) | 0.884 |
| Model 3 | 0.93 (0.71-1.22) | 0.609 | 1.28 (0.78-2.11) | 0.334 | 0.98 (0.71-1.35) | 0.895 |
| Male patients |  |  |  |  |  |  |
| Model 1 | 0.90 (0.63-1.30) | 0.576 | 1.08 (0.59-1.99) | 0.803 | 1.01 (0.65-1.57) | 0.968 |
| Model 2 | 0.92 (0.64-1.33) | 0.660 | 1.19 (0.64-2.21) | 0.586 | 1.01 (0.65-1.58) | 0.954 |
| Model 3 | 0.92 (0.64-1.33) | 0.670 | 1.20 (0.65-2.23) | 0.562 | 1.01 (0.65-1.58) | 0.956 |
| Female patients |  |  |  |  |  |  |
| Model 1 | 1.05 (0.70-1.58) | 0.803 | 1.72 (0.74-3.99) | 0.203 | 1.03 (0.64-1.65) | 0.911 |
| Model 2 | 0.95 (0.63-1.43) | 0.813 | 1.53 (0.65-3.59) | 0.327 | 0.94 (0.58-1.52) | 0.797 |
| Model 3 | 0.95 (0.63-1.43) | 0.802 | 1.50 (0.64-3.53) | 0.347 | 0.94 (0.58-1.51) | 0.791 |

CI, confidence interval; OD: odds ratio.
Model 1: adjusted for age and sex
Model 2: adjusted for age, sex, hypertension, obesity, diabetes, sleep apnoea, atrial flutter and fibrillation
Model 3: adjusted for factors in Model 2 plus irritable bowel syndrome, Crohn's disease and ulcerative colitis

## Notes

### Competing Interest Statement

The authors have declared no competing interest.

### Funding Statement

No external funding was received for this study

### Author Declarations

This study was approved by La Trobe University Human Research Ethics Committee (approval number, HEC20380).

